# Central role of the glutamate metabolism in long-term antiretroviral treated HIV-infected individuals with metabolic syndrome: a cross-sectional cohort study

**DOI:** 10.1101/2021.04.01.21254778

**Authors:** Marco Gelpi, Flora Mikaeloff, Andreas Dehlbæk Knudsen, Rui Benfeitas, Shuba Krishnan, Julie Høgh, Daniel D. Murray, Henrik Ullum, Ujjwal Neogi, Susanne Dam Nielsen

## Abstract

**Background:** Metabolic syndrome (MetS) is one of the major factors for cardiometabolic comorbidities in people living with HIV (PLWH). The long-term consequences of HIV-infection and combination antiretroviral therapy (cART) in metabolic reprogramming are unknown. In this study, we aim to investigate metabolic alterations in long-term well-treated PLWH with MetS to identify the potential mechanism behind the MetS phenotype using advanced statistical and machine learning algorithms.

**Methods:** We included 200 PLWH ≥40 years old from the Copenhagen Comorbidity in HIV-infection (COCOMO) study. PLWH were grouped into PLWH with MetS (n=100) and without MetS (n=100). The clinical data were collected from the COCOMO database and untargeted plasma metabolomics was performed using ultra-high-performance liquid chromatography/mass spectrometry (UHPLC/MS/MS). Both clinical characteristics and plasma samples were collected at study baseline. We applied several conventional approaches, machine learning algorithm and linear classification model to identify the biologically relevant metabolites associated with MetS in PLWH.

**Findings:** A total of 877 characterized biochemicals were identified. Of these, 9% (76/877) biochemicals differed significantly between PLWH with and without MetS (false discovery rate <0.05). The majority belonged to the amino acid metabolism (n=33, 43%). A consensus identification by combining supervised and unsupervised methods indicates 11 biomarkers of MetS phenotype in PLWH, of which seven (63%) have higher abundance in PLWH with MetS compared to the PLWH without MetS. A weighted co-expression network by Leiden partitioning analysis identified seven communities of positively intercorrelated metabolites, of which a single community contained six of the potential biomarkers mainly related to glutamate metabolism (glutamate, 4-hydroxyglutamate, α-ketoglutamate and γ-glutamylglutamate).

**Interpretation:** Altered amino acid metabolism is a central characteristic of PLWH with MetS and a potential central role for glutamate metabolism in establishing this phenotype is suggested.

**Funding:** Rigshospitalet Research Council, Danish National Research Foundation (DNRF126) NovoNordisk Foundation, the Swedish Research Council (2017-01330 and 2018-06156)

## Introduction

With the introduction of combination antiretroviral treatment (cART), people living with HIV (PLWH) have experienced a dramatic increase in life expectancy, with a concomitant decline in AIDS-defining morbidity and mortality^1^. A simultaneous increase in non-AIDS-associated comorbidities has been described, with a particular increment in cardiometabolic diseases, which is now one of the leading causes of death in well-treated PLWH.^2,3^ The increased risk of metabolic syndrome (MetS) associated with HIV-infection^4^ is well documented that adversely affects the cardiovascular risk profile due to MetS. However, there are still numerous gaps in understanding the pathogenesis of MetS in PLWH. Both HIV-specific (cART, ongoing HIV replication, altered gut microbiota, and immunodeficiency)^5^ and non-HIV specific factors (e.g. lifestyle)^4,5^ have been suggested to be involved in the development of MetS in the context of HIV-infection, but its exact pathogenesis remains elusive.

With the advancement of technology, high-throughput untargeted metabolomics became attractive as a novel tool to identify large numbers of metabolites and investigate molecular mechanisms of disease phenotypes not usually included in routine biochemistry analyses.^6^ The use of global untargeted metabolomics investigation had a central role in identifying novel biomarkers and potential therapeutic targets in different conditions, including type 2 diabetes and obesity.^7-9^ However, no studies are available investigating alterations in the metabolome associated with MetS in the context of HIV-infection.

Here, we aimed to identify a metabolomic signature of MetS in the context of HIV-infection to reveal key aspects of the underlying pathophysiology, which may help to improve the understanding of the pathogenesis of this disease and potentially identity new targets for the treatment of this condition. To our knowledge, this is the first study to apply several conventional and advanced bioinformatics algorithms to analyze the metabolic alterations associated with MetS in PLWH using untargeted metabolomics.

## Methods

### Study population

PLWH were recruited from the Copenhagen Comorbidity in HIV-infection (COCOMO) study, an ongoing longitudinal, observational study to assess the burden of non-AIDS comorbidities in HIV-infection. Of the 1099 participants in the COCOMO study, 100 PLWH with MetS were randomly selected and were matched to 100 PLWH without MetS. Individuals were matched according to age, sex, duration of cART, smoking status, and current CD4^+^ T-cells count. Inclusion criteria were ≥40 years of age and available measurements of the metabolome. All plasma samples were collected at study baseline concomitant with clinical assessment. Twenty uninfected controls from the background population were also included in this study to define the level of metabolites considered as normal range. Procedures for recruitment and data collection for COCOMO have been described elsewhere.^10^ Ethical approval was obtained by the Regional Ethics Committee of Copenhagen (COCOMO: H-15017350). Written informed consent was obtained from all participants.

### Clinical and biochemical assessments

Structured questionnaires were used in COCOMO to collect information about demographics, physical activity, smoking, lipid-lowering, and antihypertensive therapy.^10^ Data regarding HIV-infection were obtained from a review of medical charts.^10^ All physical examinations were performed by trained clinic staff, as previously described.^10^ Height, weight, hip, and waist measurements and body mass index (BMI) calculations were performed according to WHO guidelines. Blood pressure (BP) was measured on the left arm after 5 minutes rest with the subject in sitting position, using an automatic Digital Blood Pressure Monitor. Non-fasting venous blood was collected and analyzed for LDL-C, total cholesterol, HbA1c, and glucose. Blood samples from both COCOMO and CGPS participants were analyzed at Herlev Hospital, Copenhagen. Metabolic syndrome was defined as ≥3 of the following: (1) waist circumference ≥94 cm in men and ≥80 cm in women, (2) systolic blood pressure ≥130 mm Hg and/or diastolic blood pressure ≥85 mm Hg and/or antihypertensive treatment, (3) non-fasting plasma triglyceride level ≥1·693 mmol/L, (4) HDL level ≤1·036 mmol/L in men or ≤0.295 mmol/L in women, and (5) self-reported diabetes and/or antidiabetic treatment and/or plasma glucose level ≥11·1 mmol/L.

### Sample Preparation and untargeted metabolomics

Untargeted metabolite profiling was carried out by Metabolon Inc. (Durham, NC, USA) using ultra-high-performance liquid chromatography/mass spectrometry/mass spectrometry (UHPLC/MS/MS) as described earlier.^11,12^ Data was normalized to sample volume, log-normalized, and minimum-imputed as given by the proprietary pipeline of the provider. The metabolomics method is ISO 9001:2015 certified and the lab is accredited by the College of American Pathologists (CAP), USA.

### Statistics

Median-centered data was used for all the analyses. To verify matching between PLWH without MetS and PLWH with MetS clinical parameters, Welch’s T-test and Mann-Whitney U test were used to compare normally distributed and non-normally distributed continuous variables, respectively. Chi-Square Test was used to compare discrete variables if expected values of the contingency table were five or more. Otherwise, Fisher’s Exact Test was used. Data were tested for normality using Anderson-Darling normality test and histograms with normal distribution superimposed. As data failed the normality test (p-value<0·05), Mann Whitney U test was carried out using R to identify metabolites differing between PLHW without MetS and PLHW with MetS. Adjustment for multiple testing was performed by considering false discovery rate (FDR) <0·05.

### Bioinformatics analysis

To define metabolic alterations of the MetS in PLWH, we used a consensus of four different algorithms as the predictive signatures of MetS in PLWH. We used two conventional approaches LIMMA and Mann-Whitney U test., and one machine learning algorithm [random forest (RF)], and one linear classification model (PLS-DA) to identify the most important metabolites associated with MetS in PLWH using R packages ropls and randomForest, respectively.^13^ Both the models separate the groups, based on the importance of metabolites. The top 30 most important variables from the PLS-DA were extracted using VipPlot.^14^ For random forest, feature selection was performed using the R package Boruta. A random forest model using 10-cross fold validation was built using selected metabolites. Performance comparison of models was done using confusion matrix, ROC curves and area under the curve (AUC). As a sensitivity analysis, a similar analysis was performed using only male patients. To identify the consensus biomarkers, overlap of methodologies was represented as a Venn diagram using the R package eulerr ing ggplot2.

### Identification of mechanistic pathways

To highlight the most enriched pathways in PLHW with MetS group compared to PLHW without MetS, metabolites with differential abundance (LIMMA, FDR<0.05) were submitted to Ingenuity Pathway Analysis software (IPA) (Qiagen, US). The top 13 enriched pathways and associated metabolites were extracted, represented as a Sankey plot using R package ggalluvial and network using Cytoscape ver 3·6·1.^15^ Metabolite-metabolite interactions with high confidence (confidence > 0.7) were retrieved from databases and experiments uploaded to STITCH(v5.0) (http://stitch.embl.de/).

Association analysis was performed using R and python 3. First, metabolites with low variance were removed. Then, pairwise Spearman correlations between all metabolites were performed. Only correlations with FDR<0.05 were conserved. The distribution of correlation coefficients was plotted to evaluate the size and connection strength of the networks. Accordingly, a weighted co-expression network for significant positive associations for all samples was constructed using the python module igraph (https://igraph.org/python/). Random networks with the same dimensions were also constructed and network properties analyzed. Leiden algorithm was used for partitioning the network using the python module leidenalg.^16^ The average degree and clustering coefficient were computed for all communities and functional enrichment was performed for more than 30 metabolites. Metabolite set enrichment analysis (MSEA) was performed on these communities using gseapy first with an in-house script containing Metabolon terms then KEGG terms. Communities partition and network were exported from python and imported to Cytoscape. Metabolites with significant differential abundance between PLWH without MetS and PLWH with MetS based on LIMMA, as well as biomarkers identified by the four methodologies were highlighted in the global network.

## Results

### Clinical Characteristics

A total of 200 PLWH and 20 uninfected controls were included in the present study. PLWH was stratified according to the presence of MetS syndrome (100 individuals with and 100 individuals without MetS). Clinical characteristics of PLWH are summarized in Table 1. Briefly, no difference in age (52 (48-61) vs 52 (47-62)) and sex (male, 90 (90%) vs 90 (90%)) was found between PLWH with and without MetS. All individuals were currently on cART. Similar duration of HIV-infection (with MetS 17.2 (9.5) vs without MetS 16 (8.1) years, p-value 0.329) and cART exposure (with MetS 13.4 (7) vs without MetS 12.7 (6.1) years, p-value 0.455) was found between the groups. Larger prevalence of CD4 nadir <200 cells was found among PLWH with MetS (53 (53%) vs 38 (38%), p-value 0.039).

**Table 1.** Clinical and demographic characteristics

|  | PLWH with MetS | PLWH without MetS | P-values |
| --- | --- | --- | --- |
| **N** | 100 | 100 |  |
| **Age** | 52 (48-61) | 52 (47-62) | 0.805* |
| **Gender, Male, n (%)** | 90 (90) | 90 (90) | 1** |
| **Mode of transmission, n (%)** |  |  | 0.745** |
| Homosexual/bisexual | 73 (73) | 71 (71) |  |
| Blood transfusion | 1 (1) | 0 |  |
| PWID | 0 | 1 (1) |  |
| Heterosexual | 20 (20) | 23 (23) |  |
| Other/unknown | 4 (4) | 5 (5) |  |
| **CD4 at ART Initiation, cells/μl, median (IQR)** | 200 (82-340) | 280 (168-354) | 0.182* |
| **CD4 at sampling, cells/μl, median (IQR)** | 691 (538-865) | 700 (547-892) | 0.865* |
| **CD8 at sampling, cells/μl, median (IQR)** | 830 (620-1215) | 755 (580-970) | 0.152* |
| **Viral load (<50 copies/mL), N (%)** | 94 (94) | 98 (98) | 0.127** |
| **Duration of treatment in years, mean (sd)** | 13.4 (7) | 12.7 (6.1) | 0.455* |
| **CD4 nadir < 200 cells, n (%)** | 53 (53.5) | 38 (38.0) | 0.039** |
| **Time since HIV diagnosis, years, mean (sd)** | 17.2 (9.5) | 16 (8.1) | 0.329*** |
| **Current cART, yes, n (%)** | 100 (100) | 100 (100) | 1** |
| **BMI, mean (sd)** | 26 (3) | 23 (4) | <0.001*** |
| **Hdl, mmol/l, median (IQR)** | 0.9 (0.8-1.1) | 1.3 (1.1-1.6) | <0.001* |
| **Tgl, mmol/l, median (IQR)** | 2.7 (2.1-3.7) | 1.3 (1.1-1.7) | <0.001* |
| **Waist, cm, median (IQR)** | 102.0 (95.0-105.2) | 89.5 (85.0-97.0) | <0.001* |
| **Systolic BP, mmHg, median (IQR)** | 138.5 (131.0-145.2) | 124.5 (117.8-142.0) | <0.001* |
| **Diastolic BP, mmHg, median (IQR)** | 86.0 (79.0-91.2) | 79.0 (73.0-85.2) | <0.001* |
| **Glucose, mmol/l, median (IQR)** | 5.1 (4.6-5.7) | 5.0 (4.5-5.5) | 0.298* |

\*Mann-Whitney U test, \*\*Chi-square test, \*\*\*T-test
Abbreviations: PLWH, people living with HIV; MetS, metabolic syndrome; PWID, people who inject

### Impaired amino acid metabolism in PLWH with MetS

The untargeted metabolomics identified 877 characterized biochemicals linked mainly with lipid metabolism (n=285, 32%), xenobiotics including food components (n=223, 25%) and amino acid metabolism (n=220, 25%) (Fig 1a). The relative standard deviation (RSD) for the internal standards for the process variability was 5%. We observed that a total of 76 biochemicals differed significantly between PLWH with and without MetS (Mann-Whitney U, FDR <0.05) of which the majority were amino acids (n=33, 43%) (Fig 1a). We next used metabolite set enrichment analysis (MSEA) and network analysis to identify MetS related mechanisms in PLWH and key biochemicals. The MSEA (FDR <0.1 KEGG and HMDB) identified several affected amino acid metabolic pathways including methionine degradation, valine degradation, tyrosine degradation along the sirtuin signaling pathway that are linked to age-related diseases (Fig 1b). The network analysis identified two key molecules that interact with other metabolites, glutamate, and α-ketoglutarate, that were linked with most of the altered pathways (Fig 1c).

**Figure 1:**
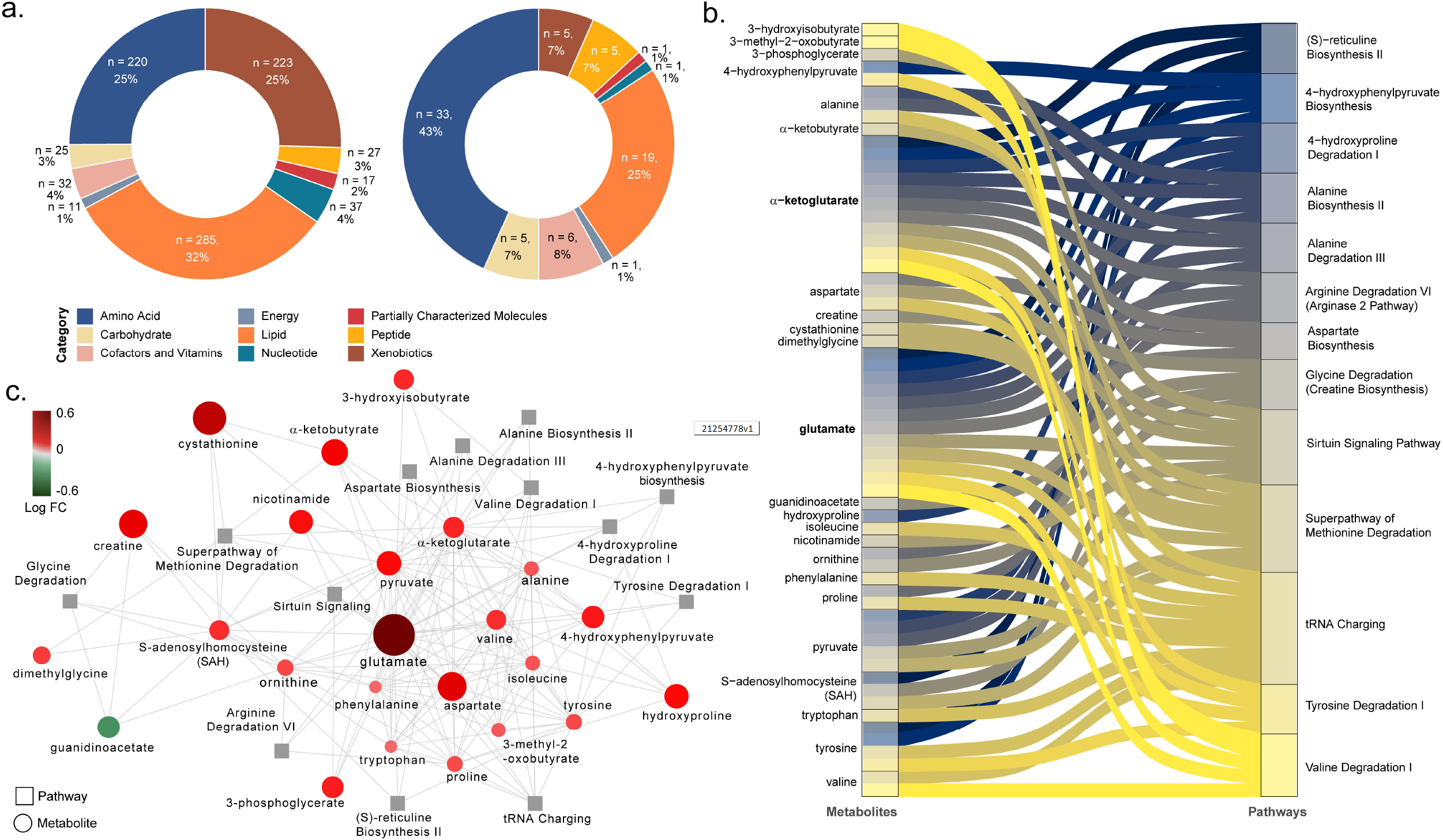
Differing metabolites and pathways found between PLWH and PLWH with MetS. (a) Doughnut charts of metabolite proportions for each super pathway for all detected metabolites (left) and metabolites with differential abundance between PLWH and PLWH with MetS (LIMMA, FDR < 0.1, n = 69). (b) Metabolites contribution to the flow of top 13 pathways represented as Sankey Plot. (c) Cytoscape network of top 13 pathways and associated enriched metabolites.

### A combination of methodologies emphasizes the role of glutamate metabolism in MetS among PLWH

By combining supervised and unsupervised methods, we sought to identify the most conservative and consensus set of metabolites associated with MetS in PLWH. Then, random forest was run on data with two classes: PLWH with and without MetS. The top-ranking metabolites (n=21, using the feature selection by Boruta) (Fig 2a) showed several metabolites such as 4-hydroxyglutamate, γ-glutamylglutarate, glutamate, α-ketoglutarate, γ-glutamylglycine and N-acetylglutamate that highlights the central role of glutamate metabolism in MetS. This model achieved 80% predictive accuracy, sensitivity and specificity of 0.8 (Figure S1), and an AUROC of 0.88 (Fig 2b). PLS-DA was also used to classify the whole data set (N = 877) that was submitted after scaling. A model using the two first components have significant coefficient of determination (R2=0.58) and coefficient of prediction (Q2=0.2). Using the predictive Variable Importance in Projection (VIP) vector, the top 30 crucial metabolites for this model were extracted (Fig S2). LIMMA identified 69 metabolites that were significantly different between the groups (FDR <0.05). A consensus identification among the four methods identified 11 biomarkers that were robustly identified by all approaches (Fig 3c and Fig S3). A UMAP projection of samples based on these 11 biomarkers showed a clear separation of the uninfected controls and PLWH with MetS (Fig 2d). Seven of our potential biomarkers were upregulated in PLWH with MetS: 1-carboxyethylleucine, 4-cholesten-3-one, 4-hydroxyglutamate, α-ketoglutamate, γ-glutamylglutamate, glutamate, and isoleucine, while four were downregulated: carotene diol(2), glycerate, PSP and PC/3-MAPC (Fig 2e). γ-glutamylglycine and orotate were found only in RF while N2,N5-diacetylornithine, γ-glutamyl-alpha-lysine and γ-glutamyltyrosine were identified only by PLS-DA. Though our study population was highly matched, majority of the PLWH were male and MetS categorizations varied between male and female. We also perform sensitivity analysis restricted only to the males. Among the metabolites, 13 were identified as consensus between all the analysis (Fig S4a). The RF model showed slightly lower performance than the full model (AUROC=0·85, Fig S4b). Interestingly, among the 13 potential biomarkers identified by the same process in males, eight were shared with biomarkers identified in the entire sample population and are mainly linked with glutamate metabolism (Fig S4c). The five other biomarkers of MetS in males were γ-glutamylvaline, N-acetylglutamate, N-acetylvaline, γ-glutamylisoleucine and valine (Fig S4d) with an increase in PLWH with MetS indicating the altered glutamate metabolism as a common feature in PLWH with MetS. Despite metabolic changes associated with MetS, confirmation that our potential biomarkers were specific for PLWH with MetS and not biomarkers of obesity or treatment was necessary.

**Figure 2:**
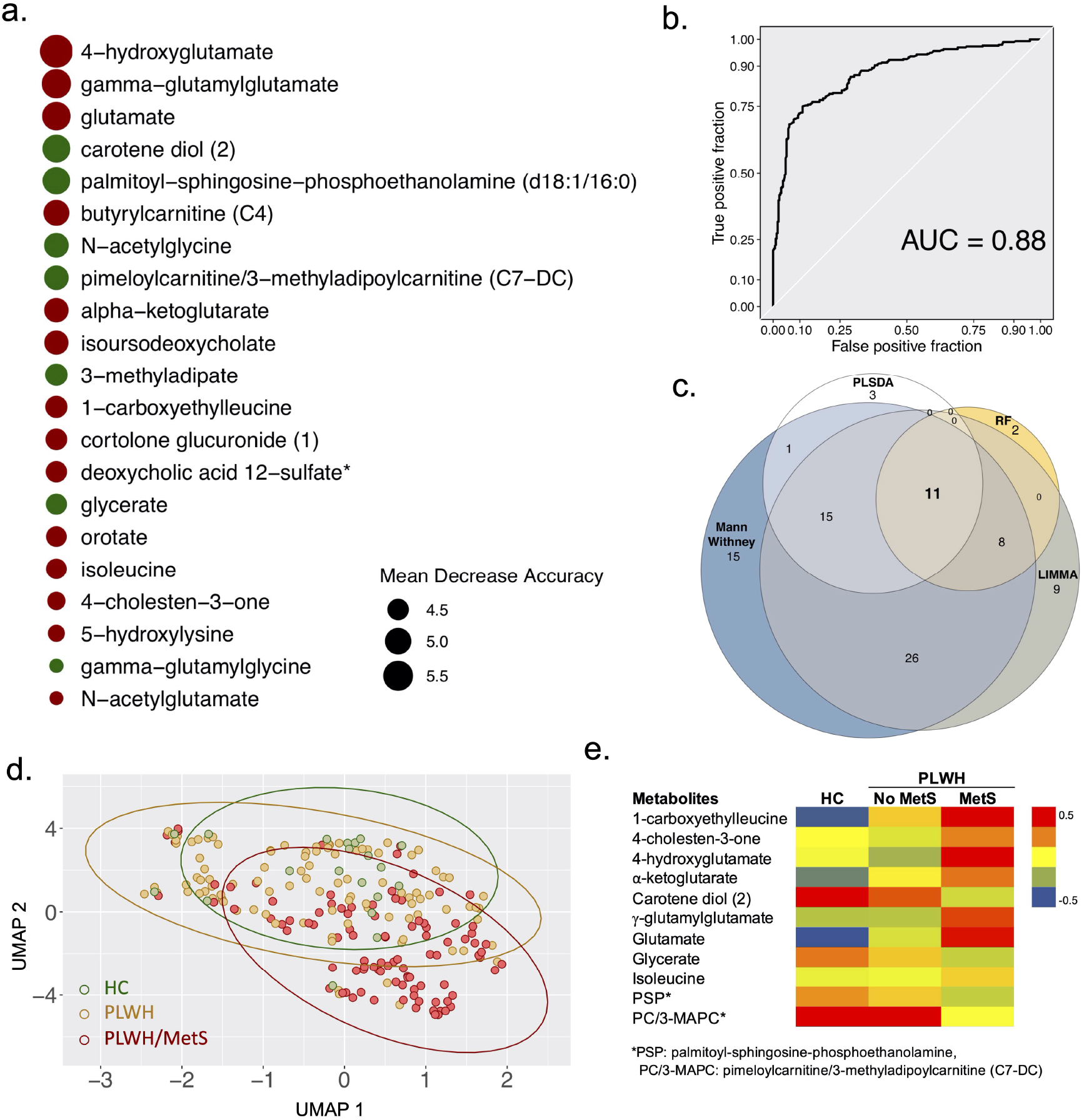
Biomarkers (n=11) with differential abundance between PLWH and PLWH with MetS identified by four methodologies. (a) Bubble plot representing Random forest variable importance based on mean decrease accuracy (a measure of the model’s performance without each metabolite). Values are scaled by the standard error of the measure. Metabolites represented at the top of the figure are the most important for prediction. (b) Receiver Operating Characteristic (ROC) curve of random forest classifier. (c) Venn diagram summarizing biomarkers identified by Mann-Whitney U test, LIMMA, Random Forest (RF), and PLS-DA. (d) UMAP visualization of the 11 biomarkers. Controls (green) and PLWH (yellow) are segregating from PLWH with MetS (red). (e) Heatmap showing log2 intensities of the 11 biomarkers in HC, PLWH without MetS and PLWH with MetS.

**Figure 3:**
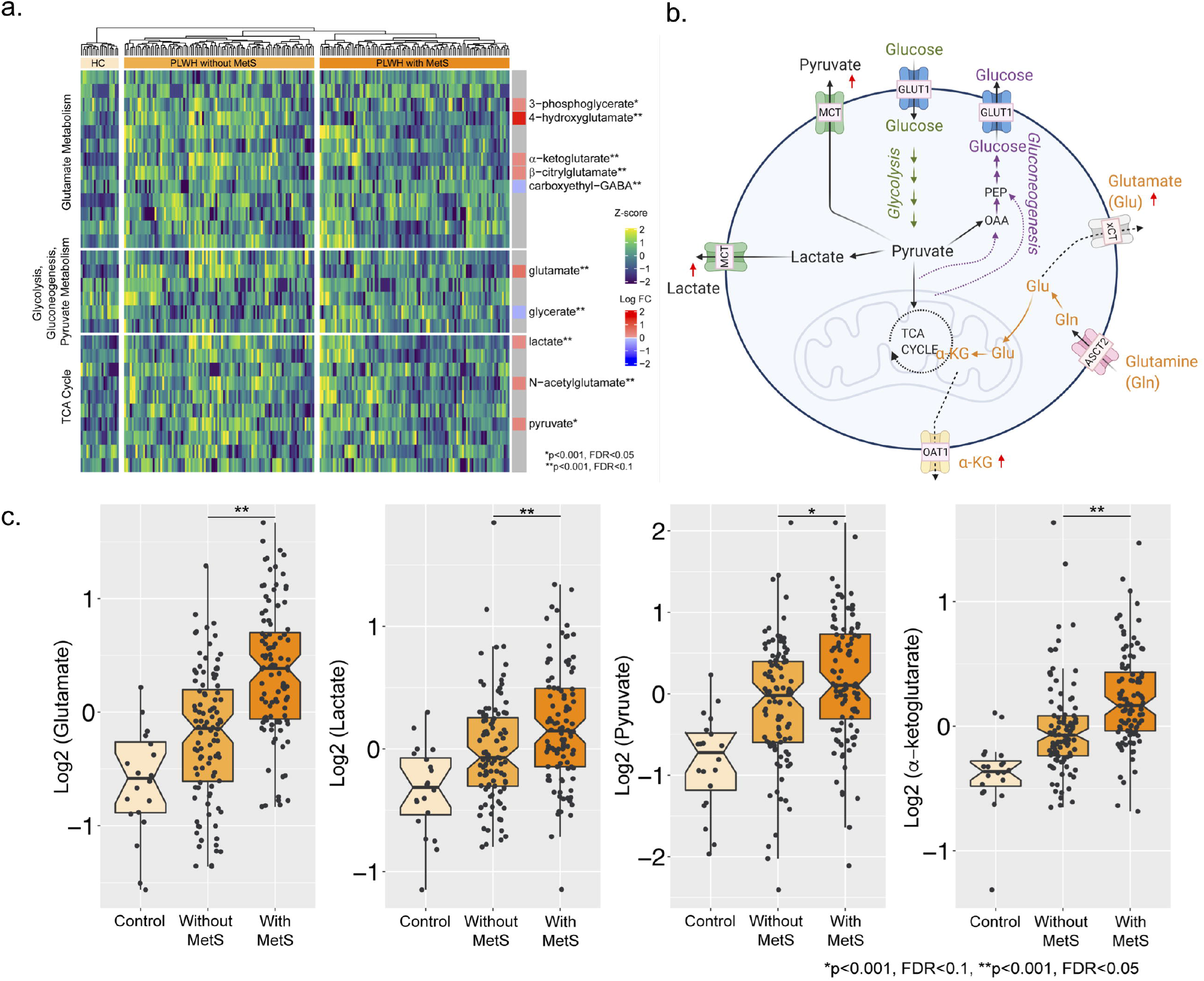
Central carbon metabolism with higher efflux of key metabolites. (a) Heatmap showing the level of metabolites of glutamate metabolism, glycolysis/gluconeogenesis/pyruvate metabolism, and TCA cycle. The statistically significant differentially abundant metabolites are marked with single asterisk at the level of p <0.001 and FDR <0.1 and double asterisk p <0.001 and FDR <0.05 using LIMMA. (b) Schematic representation of the central carbon metabolism with the key pathways. A red up arrow indicates an increase in plasma of PLWH with MetS compared to the PLWH. (c) level of key metabolites alpha-ketoglutarate and glutamate of glutamate metabolism and major carbohydrates lactate and pyruvate. Box plots showing log2 intensities of Controls, PLWH without MetS and PLWH with MetS. Single asterisks indicate statistically significant differences p<0·001 and FDR<0·1 and double asterisk p<0.001 and FDR<0·05.

We thus tested whether there was an association between biomarker and MetS status corrected for BMI. All biomarkers were significant (FDR<0.05) after BMI and treatment correction (Table S1, Table S2). Pharmaceuticals can also affect metabolic profiles. Five xenobiotics were found to be significantly different between PLWH with and without MetS: hydrocinnamate, abacavir, losartan, tartronate and thioproline. Correction for xenobiotics level was performed for each metabolite using linear regression (Table S3). All metabolites passed the correction except glycerate. After correlation analysis, glycerate was shown to have significant correlation with tartronate (R=0.8, FDR<0·00001) (Fig S5).

### Impaired central carbon metabolism with higher efflux of key carbohydrates

Higher levels of plasma glutamate and altered glutamate metabolism may indicate dysregulation of central carbon metabolism. Therefore, we investigated key metabolites from other pathways of central carbon metabolism: glycolysis, gluconeogenesis, and TCA cycle (Fig 3a). No changes were observed in plasma glucose levels (as shown in Table 1 also, glucose being part of routine blood biochemistry), but we found a significant increase in pyruvate, lactate, and α-ketoglutarate levels in PLWH with MetS compared to PLWH without MetS (Fig 3b) Apart from the higher abundance in the PLWH with MetS, the median (IQR) of glutamate, lactate, pyruvate and a-ketoglutarate metabolites were also statistically higher in PLWH compared to uninfected controls (FDR<0.05) (Fig 3c). This further indicates the increased efflux of glycolytic and TCA cycle metabolites from cells into the bloodstream in PLWH.

### Community analyses confirm identified biomarkers and display possible mechanisms of metabolic syndrome in PLWH

To establish the metabolic shifts (i.e., the metabolic alterations due to disease conditions) associated with MetS in PLWH, a weighted co-expression network analysis was performed using all detected metabolites and all patients. In this network, the nodes were individual metabolites, and edges were modeled by the positive coefficients of correlation between two metabolites (Spearman, FDR < 0·05). The resulting network consisted of 867 nodes and 45379 edges. Leiden partitioning highlights seven communities of positively intercorrelated metabolites (Fig 4 top). Analysis at pathway level revealed that the most central community, represented as the center of the network, was enriched for the KEGG terms 2-oxocarboxylic acid metabolism, organic acids, and lipids (Metabolon− category). This community displayed a very heterogeneous pattern, where only six differentially expressed metabolites (out of 73) were present and half had higher abundance in PLWH with MetS. Interestingly, we observed that a single community contained six potential biomarkers previously identified (Fig 4 bottom). This community also displayed 38 differentially expressed metabolites, of which only one was lower abundance in PLWH with MetS and contained six of the 11 potential biomarkers identified above. These biomarkers showed many interconnections between each other, though other biomarkers including 4-cholesten-3-one were segregated in other communities. This community was enriched in common amino acids (KEGG and Metabolon) and peptides (Metabolon).

**Figure 4:**
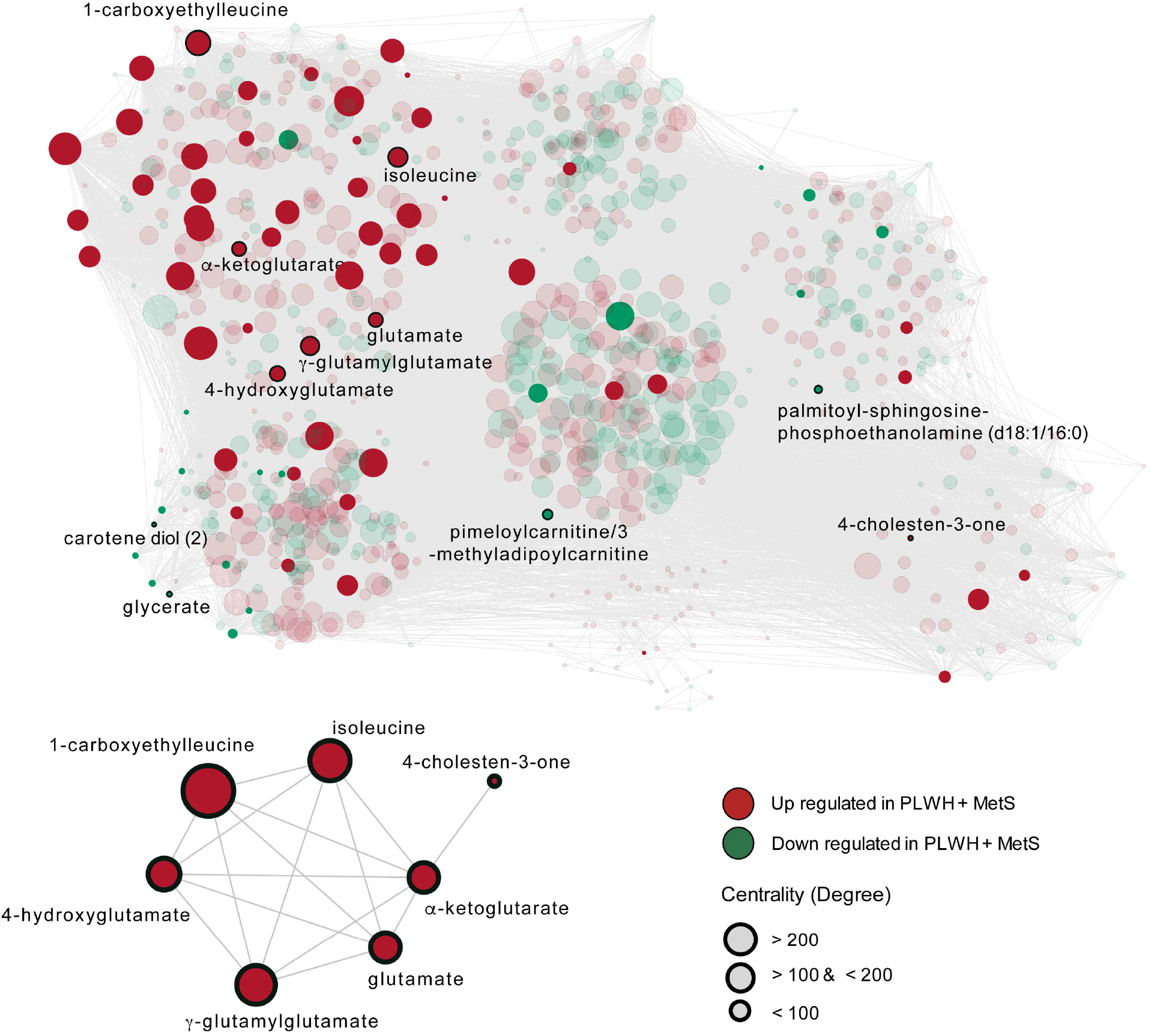
The metabolome-wide weighted co-expression network. A weighted metabolite co-expression network was generated (positive Spearman rank correlations, FDR<0·05). Significant metabolites based on LIMMA have represented opaques and non-significant transparent. Biomarkers found in the first community are represented (bottom-left).

## Discussion

The present study resulted in two key findings regarding MetS in the context of well-treated HIV-infection. Using a consensus approach of traditional biostatistics and advanced machine learning algorithms, we first identified altered amino acid metabolism as a central characteristic of PLWH with MetS with glutamate metabolism as a key metabolic pathway in this phenotype. Second, a weighted co-expression network analysis indicates the interaction between lipid metabolism and amino acid metabolism that could modulate glutamate metabolism as a coordinated metabolic alteration in PLWH with MetS.

The role of amino acid metabolism in the development of metabolic dysfunction has been extensively studied in the general population^16^, but it still unclear in the context of HIV-infection. The most consistent findings describe an essential role for the three branched-chain amino acids (BCAA) (leucine, isoleucine, and valine) in the development of cardiometabolic diseases, in particular obesity and diabetes (as reviewed in^17^). In our study, we observed higher levels of isoleucine associated with MetS among PLWH. This finding is in line with previous studies describing increased isoleucine levels in nascent metabolic syndrome in the general population.^18^ All three BCAA are catabolized into glutamate and a branched-chain-ketoacid (BCKA) via the transamination of alpha-ketoglutamate by branched-chain aminotransferase (BCAT). BCKA is further metabolized by the activity of the enzyme branched-chain-ketoacid dehydrogenase (BCKD). Alterations of BCAT and BCKD activity have been described in individuals with MetS syndrome.^19^ In particular, lower expression of BCKD was described in visceral, but not subcutaneous, adipose tissue of metabolically impaired subjects, leading to reduced activity of this metabolic pathway and consequent accumulation of BCAA and glutamate. Accordingly, glutamate concentrations have been efficiently used to differentiate individuals with an excess of adipose tissue at abdominal level and metabolic risk.^20^ Taken together these findings suggest a potential role for alterations of BCAA catabolism in abdominal adipocytes in the pathogenesis of MetS in the background population. Previous results from our group proposed excess of abdominal adipose tissue as a critical factor in the excess risk of MetS in the context of HIV compared to uninfected individuals.^4^ The identification of metabolome alterations in PLWH with MetS may help to explain this not yet wholly understood association. In the present study, increased levels of several metabolites involved in glutamate metabolism characterized PLWH with MetS. This finding may point towards a significant association between glutamate and MetS among PLWH, thus supporting previous studies in uninfected individuals.^18^ Given the previously described association of HIV-infection with alterations of both abdominal obesity^4^ and glutamate metabolism,^12,21^ one may speculate that the role of glutamate metabolism in the pathogenesis of MetS might be even more central in the context of HIV.

In the present study, PLWH both with and without MetS had higher concentrations of several metabolites involved in glutamate metabolism compared to uninfected controls.Although the sample size of uninfected controls is small, these findings support a previous study that showed higher glutamate in PLWH than uninfected controls.^21^ The association between HIV-infection and glutamate metabolism has also been studied in the context of neurologic comorbidities.^22^ Accordingly, increased release of glutamate in the extracellular space by HIV-infected macrophages mediated by the viral protein Vpr has been suggested to play a central role in the pathogenesis of HIV-mediated neurotoxicity.^23^ It is to be noted that both lactate and pyruvate were also increased in the PLWH with cART and PLWH without MetS. It is known that lactate and pyruvate reduced glutamate-induced neurotoxicities in animal experiments.^24^ One may speculate that increased lactate and pyruvate could be due to the increased glutamate thereby linked with the severity of the MetS due to impaired aerobic metabolism and elevated metabolic diseases in PLWH. However, this may have a protective effect on neurological impairment.^25^ However, whether perturbed glutamate homeostasis is also involved in other non-AIDS-associated comorbidities like MetS is still unknown. Abdominal adipose tissue accumulation, a well-known determinant of MetS in PLWH, has been previously suggested to be associated with increase macrophage infiltration and activation.^26^ It may be speculated that alterations in macrophage glutamate metabolism in the abdominal tissue are also involved in MetS in PLWH. Thus, novel treatments targeted at reducing plasma glutamate concentration could potentially be implemented along with cART to prevent both neurological and metabolic complications in PLWH with MetS and increased build-up of lactate and pyruvate, also known for toxic proprierties.^27^ A glutamatergic drug that directly modulates the excitatory glutamate in the body or brain can potentially be used after appropriate clinical studies.

The present study has several limitations. First, due to the cross-sectional design, no conclusions on causality can be drawn. Second, the relatively low number of uninfected individuals included and the lack of clinical data in this population prevented us from investigating the impact of HIV-infection on the metabolites and pathways observed to be associated with MetS. Also, the uninfected controls were used to identify the metabolites’ normal range as the untargeted metabolomics analysis dependent upon the run, thus not used for any statistical analysis. Finally, despite adjusting for multiple testing, the high number of associations tested may have led to type I errors. However, the major strength of this study is the use of relatively larger, well-characterized, clinically matched cohorts of PLWH. To the best of our knowledge this is the first study investigating metabolome alterations associated with MetS in the context of HIV-infection.

In conclusion, we presented data suggesting alterations in glutamate metabolism to be associated with MetS in PLWH. Glutamate plays a central role in multiple metabolic pathways for the interchange of amino nitrogen by both amino acid synthesis as well as degradation, and glutamate toxicity is associated with age-related neurodegenerative disorders. Further studies are warranted to address a possible direct role of glutamate in the pathogenesis of MetS and its potential role as a biomarker for accelerated cognitive and metabolic aging in the context of HIV-infection.

## Data Availability

All data available within the manuscript.

https://figshare.com/articles/dataset/_/14356754

## ACKNOWLEDGEMENTS

We thank all the study subjects for their participation. We thank the staff at the Department of Infectious Diseases at Rigshospitalet and at Hvidovre Hospital for their dedicated participation. The computations were enabled by resources in the project [Dnr. SENS2017550] provided by the Swedish National Infrastructure for Computing (SNIC) at UPPMAX, partially funded by the Swedish Research Council through grant agreement no. 2018-05973.

