## Supplemental Figure S1-S5 and Table S1-S3 for "Central role of the glutamate metabolism in long-term antiretroviral treated HIV-infected individuals with metabolic syndrome: a cross-sectional cohort study"

### Supplementary figures

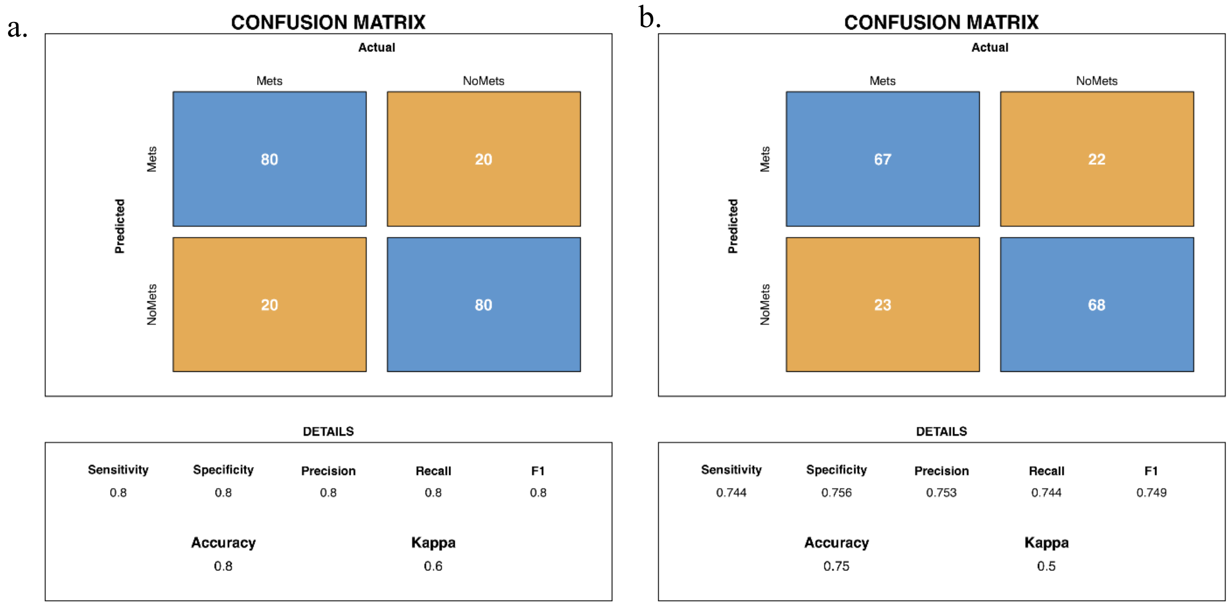


<u>Figure S1</u> : Confusion matrices for random forest models (a) complete data set (b) males only.

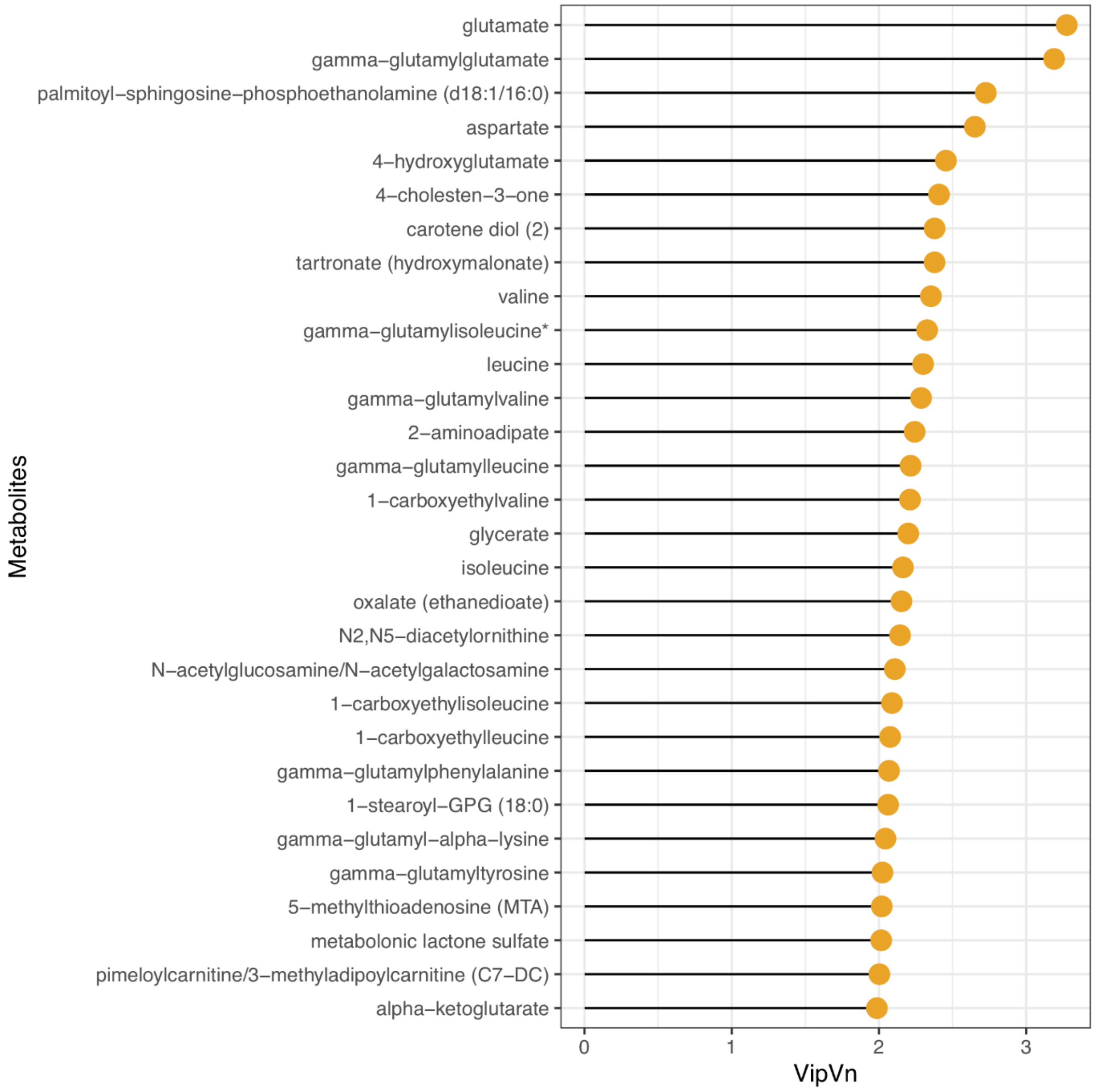


Figure S2 : Variable Importance plot representing top feature importance extracted from PLS-DA model in complete data set.

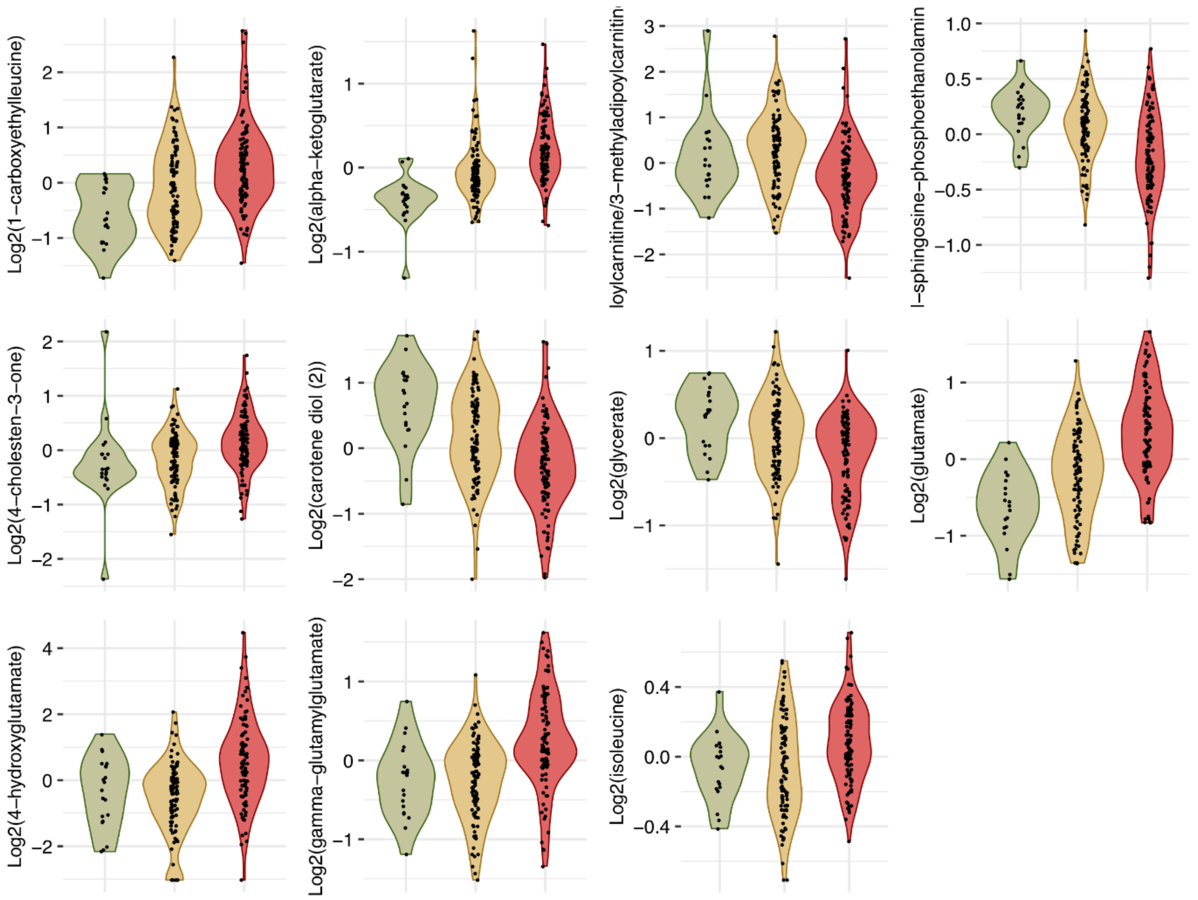


Figure S3: Violin plots showing log2 intensities of 11 identified biomarkers in Controls (green), PLWH (yellow) and PLWH+MetS (red) patients.

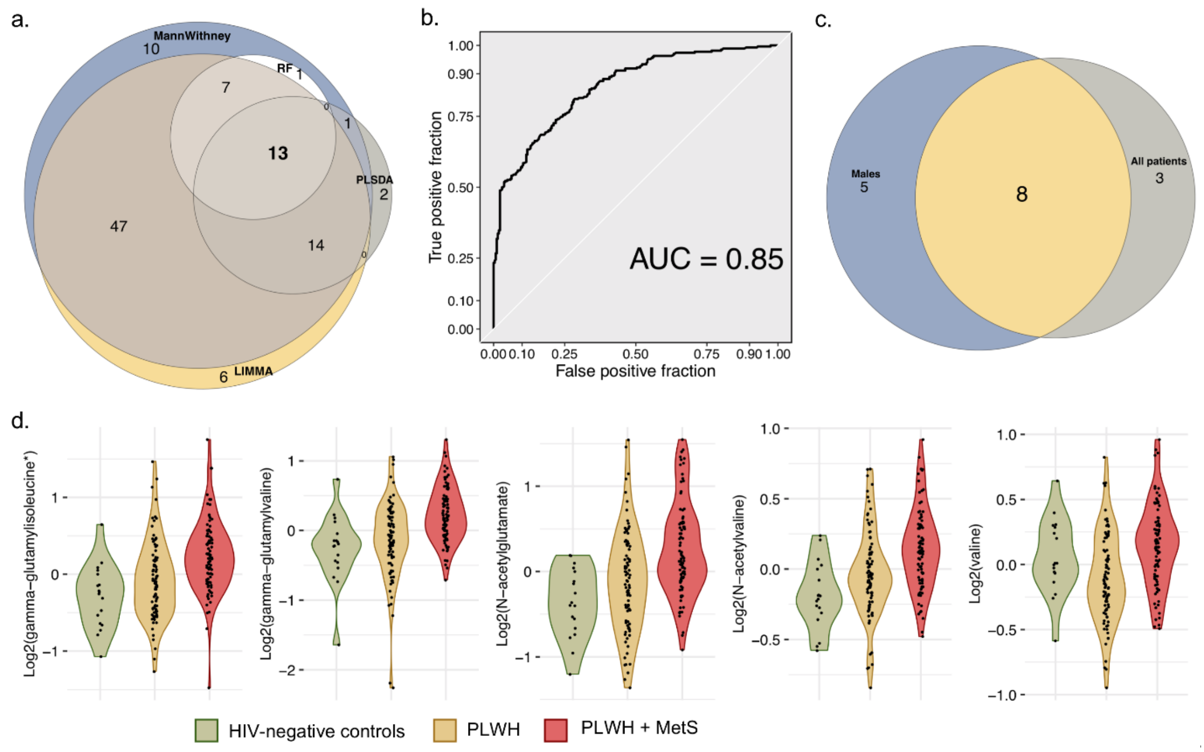


**<u>Figure S4</u>: Biomarkers with differential abundance between males PLWH and PLWH with MetS.** (a) Venn diagram summarizing biomarkers identified by Man Whitney U test, LIMMA, Random Forest (RF) and PLS-DA in male patients. (b) ROC curve of random forest classifier for predicting metabolic syndrome status in PLWH in male patients. (c) Venn diagram representing the overlap between biomarkers identified in all patients and in males. (d) Violin plots showing log2 intensities of 5 identified biomarkers in Controls (green), PLWH (yellow) and PLWH with MetS (red) specific to male patients.

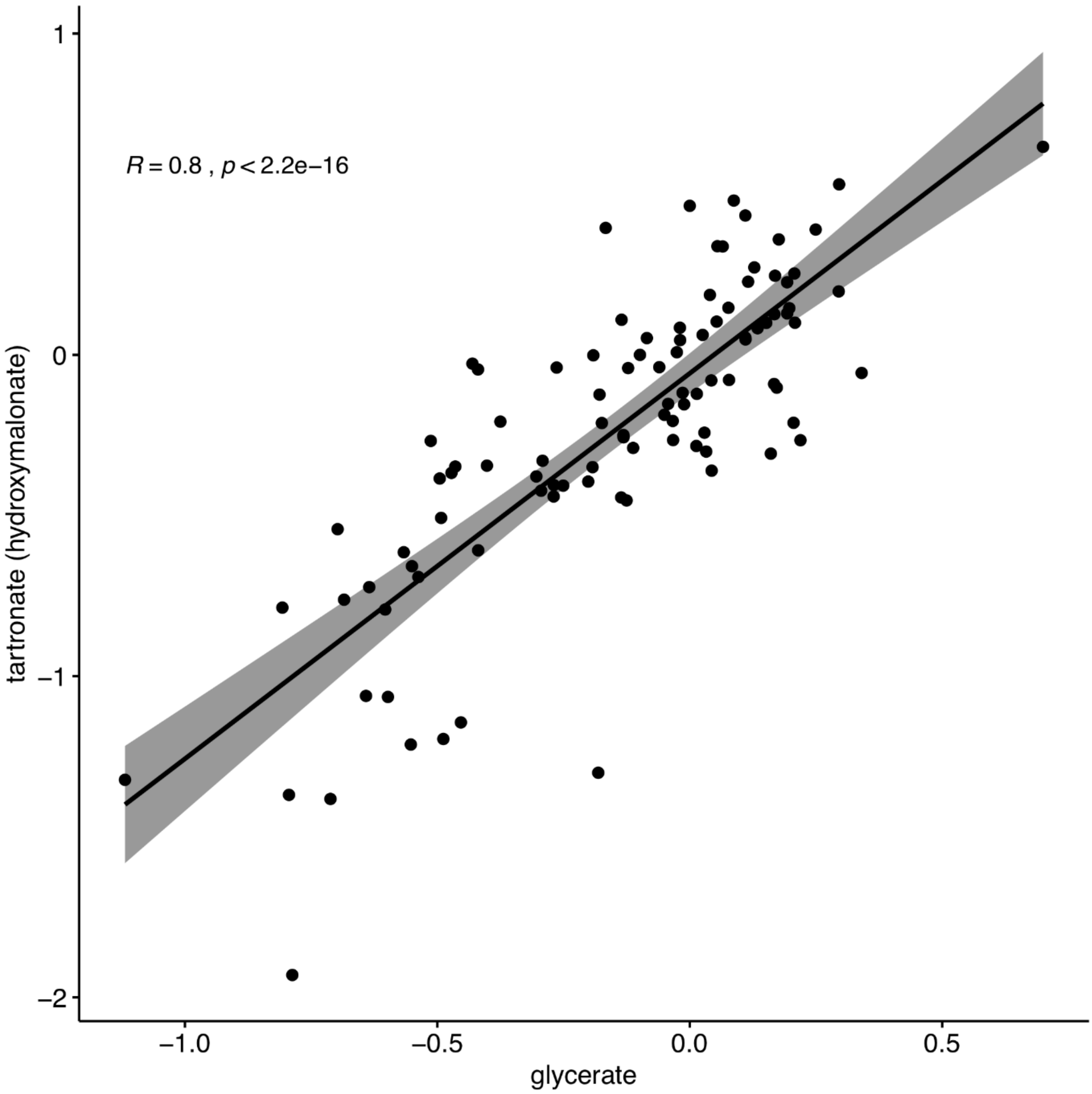


Figure S5: Scatter plot representing relationship between scaled intensities of glycerate and tartronate. The regression line, Pearson correlation coefficient and p-value are indicated.

Table S1 : Linear regression models adjusted for BMI and treatment regimen (PI, NNRTI and INSTI) showing the association of 11 biomarkers with metabolic syndrome in PLWH.

| | Estimate | StdError | tStat | pvalue | padjust |
|---|---|---|---|---|---|
| **X1.carboxyethylleucine** | 0.485 | 0.141 | 3.456 | <0.001 | 0.001 |
| **X4.cholesten.3.one** | 0.198 | 0.059 | 3.307 | 0.001 | 0.001 |
| **X4.hydroxyglutamate** | 1.412 | 0.332 | 4.252 | <0.001 | <0.001 |
| **alpha.ketoglutarate** | 0.18 | 0.053 | 3.368 | <0.001 | 0.001 |
| **carotene.diol..2.** | -0.34 | 0.081 | -4.196 | <0.001 | <0.001 |
| **gamma.glutamylglutamate** | 0.379 | 0.068 | 5.545 | <0.001 | <0.001 |
| **glutamate** | 0.393 | 0.072 | 5.386 | <0.001 | <0.001 |
| **glycerate** | -0.169 | 0.047 | -3.544 | <0.001 | <0.001 |
| **isoleucine** | 0.093 | 0.028 | 3.295 | 0.002 | 0.001 |
| **palmitoyl.sphingosine.phosphoethanolamine..d18.1.16.0.** | -0.157 | 0.036 | -4.415 | <0.001 | <0.001 |
| **pimeloylcarnitine.3.methyladipoylcarnitine..C7.DC.** | -0.396 | 0.134 | -2.962 | 0.003 | 0.003 |

Table S2. Linear regression models adjusted for BMI and treatment regimen (AZT, TDF, TAF and ABC) showing the association of 11 biomarkers with metabolic syndrome in PLWH.

| | Estimate | StdError | tStat | pvalue | P adjust |
|---|---|---|---|---|---|
| **X1.carboxyethylleucine** | 0.426 | 0.152 | 2.797 | 0.006 | 0.006 |
| **X4.cholesten.3.one** | 0.268 | 0.063 | 4.194 | <0.001 | <0.001 |
| **X4.hydroxyglutamate** | 1.332 | 0.365 | 3.650 | <0.001 | <0.001 |
| **alpha.ketoglutarate** | 0.151 | 0.058 | 2.597 | 0.010 | 0.0101775 |
| **carotene.diol..2.** | -0.339 | 0.089 | -3.777 | <0.001 | <0.001 |
| **gamma.glutamylglutamate** | 0.318 | 0.072 | 4.427 | <0.001 | <0.001 |
| **glutamate** | 0.314 | 0.077 | 4.060 | <0.001 | <0.001 |
| **glycerate** | -0.147 | 0.047 | -3.154 | 0.002 | 0.003 |
| **isoleucine** | 0.089 | 0.030 | 2.947 | 0.004 | 0.004 |
| **palmitoyl.sphingosine.phosphoethanolamine..d18.1.16.0.** | -0.152 | 0.038 | -3.948 | <0.001 | <0.001 |
| **pimeloylcarnitine.3.methyladipoylcarnitine..C7.DC.** | -0.449 | 0.145 | -3.082 | 0.002 | 0.003 |

Table S3: Linear regression models adjusted for xenobiotics levels showing the association of 10 biomarkers with metabolic syndrome in PLWH.

| | Estimate | StdError | tStat | pvalue | padjust |
|---|---|---|---|---|---|
| **1-carboxyethylleucine** | 0.274 | 0.145 | 1.878 | 0.062 | 0.068 |
| **4-cholesten-3-one** | 0.533 | 0.144 | 3.687 | <0.001 | <0.001 |
| **4-hydroxyglutamate** | 0.533 | 0.143 | 3.723 | <0.001 | <0.001 |
| **alpha-ketoglutarate** | 0.447 | 0.148 | 3.025 | 0.003 | 0.003 |
| **carotene diol (2)** | -0.459 | 0.144 | -3.191 | 0.002 | 0.003 |
| **gamma-glutamylglutamate** | 0.719 | 0.133 | 5.376 | <0.001 | <0.001 |
| **glutamate** | 0.743 | 0.134 | 5.511 | <0.001 | <0.001 |
| **glycerate** | -0.036 | 0.088 | -0.412 | 0.680 | 0.681 |
| **isoleucine** | 0.413 | 0.146 | 2.816 | 0.005 | 0.007 |
| **palmitoyl-sphingosine-phosphoethanolamine (d18:1/16:0)** | -0.669 | 0.141 | -4.739 | <0.001 | <0.001 |
| **pimeloylcarnitine/3-methyladipoylcarnitine (C7-DC)** | -0.469 | 0.149 | -3.147 | 0.002 | 0.003 |
